# Real-World Marijuana Use Is Associated With Altered Nocturnal Cardiac Autonomic Function, Reduced Sleep Quality, and Declining Consumption During Wearable Monitoring

**DOI:** 10.64898/2026.09.15.26363138

**Authors:** Dylan Curran, William von Hippel, Finnbarr Fielding, Summer R. Jasinski, Josh Leota, Jenna G. Cohen, Kristen E. Holmes, Gregory J. Grosicki, Christian P. Cheung

**Author notes:** Co-Senior Authors (Gregory J. Grosicki, Christian P. Cheung). **Corresponding Author:** Christian P. Cheung, WHOOP, Inc., One Kenmore Square, Boston, USA, 02215.

## Abstract

**Background:** Marijuana is among the most widely used controlled substances, with use rising as risk perception declines. Despite growing acceptance, concerns remain regarding its effects on cardiovascular and autonomic physiology, sleep, and risk of cannabis use disorder. Although laboratory and epidemiological studies have characterized these associations, objective evidence from real-world settings is limited.

**Methods:** We studied cardiac autonomic function, sleep, and marijuana use patterns in 16,965 consumer-wearable device users, contributing a total of 5,226,739 person-days of data with or without self-reported use of marijuana. Linear mixed models related day-level use to nocturnal cardiac autonomic function (resting heart rate [RHR], heart rate variability [HRV]) and sleep (duration, fragmentation, architecture) at the within- and between-person levels. Binomial mixed models were used to estimate use probability across 72 weeks of marijuana tracking.

**Results:** Members reported marijuana use on a median of 25.1% of logged days (IQR, 7.1–62.6%). Within individuals, use was associated with lower RHR (−0.12bpm; 99.5% CI: [−0.14, −0.10]) and lower HRV (−0.47ms [−0.53, −0.42]). Sleep was longer (+7.60min [7.37, 7.83]) but more fragmented, with more disturbances (+0.47 [0.45, 0.48]), greater wake time (+1.79min [1.70, 1.89]), and altered wearable-estimated sleep architecture. Daily use probability declined over 72 weeks (−11.1% [−11.5, −10.8]).

**Conclusions:** Marijuana use was associated with altered nocturnal cardiac autonomic physiology and longer but more disrupted sleep. Marijuana use declined after commencing tracking, raising the possibility that longitudinal self-monitoring may influence use. These findings extend laboratory evidence on marijuana use to real-world settings and demonstrate the potential of consumer wearables for studying substance use and its physiological correlates at scale.

**Highlights:**

- Marijuana-use nights were associated with lower resting heart rate and heart rate variability.
- Sleep was prolonged but more fragmented with marijuana use.
- Marijuana use fell over 72 weeks of wearable tracking.

## INTRODUCTION

Marijuana is amongst the most widely used controlled substances globally, with an estimated 244 million individuals reporting use in 2023 (*World Drug Report*, 2025). Use continues to increase alongside expanding legalization, decriminalization, access through regulated markets, and declining perceptions of risk (Manthey et al., 2021; McDonald et al., 2025; Waddell, 2022; Yu et al., 2020). These changes have been accompanied by the commercialization of marijuana and the growing promotion of products for specific health, wellness, and functional purposes (Moran et al., 2025; Nali et al., 2025). As marijuana becomes more accessible and integrated into everyday health behaviors, understanding its physiological effects under real-world conditions is increasingly important.

A central uncertainty is whether perceptions of marijuana’s physiological effects align with those measured objectively. Sleep is among the most frequently reported reasons for marijuana use (Dzierzewski et al., 2026). Users report that cannabis relaxes the mind and body and helps them achieve deeper, longer, and more uninterrupted sleep (Stueber and Cuttler, 2023), and self-report and naturalistic studies have further linked cannabis use to reduced sleep-onset latency, fewer nighttime awakenings, and relief of insomnia symptoms (Goodhines et al., 2019; Vigil et al., 2018; Winiger et al., 2021). However, these perceived benefits are not consistently supported by controlled laboratory studies using polysomnography, with recent meta-analyses suggesting no consistent effect on total sleeping time, wake after sleep onset, or sleep architecture (Velzeboer et al., 2025). Marijuana also affects cardiac autonomic and cardiovascular physiology (Benowitz and Jones, 1977, 1975; Cheung et al., 2022; D’Souza et al., 2025; Jones, 2002; Nardone et al., 2023), and observational studies have associated its use with adverse cardiovascular outcomes (Jeffers et al., 2024; Kamel et al., 2025). Whether these laboratory and epidemiological findings translate to habitual, self-directed marijuana use in everyday life remains unclear.

Consumer wearables offer an opportunity to address this gap. While laboratory studies provide important mechanistic insight and experimental control by administering fixed products, doses, and timings under artificial conditions, they may not capture how habitual marijuana use affects physiology, nor reflect the timing and dosage that people self-select in response to the events of everyday life. By combining continuous physiological measurement with daily behavioral reports, wearables can characterize associations between marijuana use, sleep, and cardiac autonomic function across the natural variation in how individuals use marijuana. This approach may help bridge perceived effects, experimental evidence, and physiological responses observed in everyday life.

Wearable devices also provide insight into longitudinal use, with prior studies utilizing wearable devices indicating declines in tobacco and alcohol use (Curran et al., 2026; G. J. Grosicki et al., 2026). These findings raise the possibility that repeated tracking, feedback, or increased awareness may similarly accompany changes in marijuana use, a particularly important area of investigation given increasing cannabinoid concentrations within marijuana products (ElSohly et al., 2016), and recent reports of a growing prevalence of marijuana use disorder (Han et al., 2026). Whether marijuana use changes over the course of longitudinal self-monitoring with a wearable platform remains unknown.

The purpose of the present study was to leverage large-scale physiological and behavioral data to examine the real-world effects of marijuana use across three domains. To do so, we assembled what is, to our knowledge, the largest real-world dataset of marijuana use, sleep, and cardiac autonomic function to date, comprising 5,226,739 person-days from 16,965 individuals. First, we compared resting heart rate (RHR) and heart rate variability (HRV) between marijuana use and non-use days to characterize differences in cardiac autonomic physiology. Second, we compared wearable-derived sleep measures between use and non-use days. Finally, we characterized marijuana use trajectories over the course of 72 weeks. We hypothesized that marijuana-use days would differ from non-use days in both cardiac autonomic and sleep metrics, and that marijuana use would decline over time.

## METHODS

### Ethics Statement

All participants consent to the use of their deidentified data for research purposes when they join the consumer wearable platform and are given the opportunity to revoke consent and have their data deleted at any time. The study protocol was reviewed and approved by an independent institutional review board (Salus Institutional Review Board; approval number 251121).

### Study Design

We conducted a retrospective observational cohort study using data collected during routine use of a consumer wearable (WHOOP, Boston, USA). The device continuously records physiological data and includes an in-app daily journal in which members can optionally report engagement in selected behaviors, including marijuana use. We examined overnight cardiac autonomic and sleep measures in relation to day-level self-reported marijuana use and characterized the daily probability of use over 72 weeks. Each member was observed for 504 days after first logging marijuana use, with observations beginning between January 2022 and June 2024. Because self-reported marijuana use varied from day to day, each member served as their own comparator across days with and without use.

### Participants

Participants were members who began logging marijuana use between January 2022 and June 2024. Eligible members were 18 to 79 years of age, self-identified gender as man or woman, and had at least three days with and without self-reported marijuana use, and at least eight weeks with an explicit yes-or-no marijuana response on all seven days. From eligible members, a random sample of up to 10,000 per gender was drawn, with the final cohort including a total of 10,000 men and 6,965 women.

### Self-Reported Marijuana Use

Daily marijuana use was self-reported through the WHOOP smartphone application via the WHOOP Journal feature, a customizable morning prompt through which users answer questions about behavior from the prior day. Users were prompted to indicate whether they “used marijuana” (“yes” or “no”). Only days marked explicitly “yes” or “no” were included.

### Wearable Device & Platform Derived Metrics

Sleep and cardiac autonomic metrics were quantified via the WHOOP wearable device via previously described photoplethysmography and accelerometry methods (Grosicki et al., 2026; Holmes et al., 2024; Sargent et al., 2024). The device detects major sleep periods of at least one hour in duration. Within each detected sleep period, each 30-second interval is classified into one of four sleep stages: wake, light, slow-wave sleep, and rapid eye movement (REM) sleep. WHOOP has been validated against electrocardiography and polysomnography for heart rate (99% agreement) and two-stage sleep categorization (86–89% agreement) (Bellenger et al., 2021; Miller et al., 2022, 2020). Sleep duration was defined as total sleep time (light, slow-wave sleep, and REM). Sleep stages were expressed both in minutes and as a percentage of total sleep duration. Sleep fragmentation was characterized via the number of sleep disturbances, wake duration, arousal time, and sleep efficiency (the percentage of time in bed spent asleep). Wake duration was defined as the total time spent awake within the sleep period, including all wakefulness from brief involuntary arousals to longer conscious awakenings, whereas arousal time captured only the cumulative duration of brief, transient arousals (short shifts toward wakefulness lasting seconds that may not reach conscious awareness) and thus represents a subset of wake duration. Throughout each sleep period, the device derives peak-to-peak heartbeat intervals from the photoplethysmography to measure RHR and estimate HRV, calculated as the root mean square of successive differences (RMSSD). To generate nightly RHR and HRV values, WHOOP first filters out epochs identified as wake or as having low signal quality. The remaining epochs are then aggregated using a weighted average, in which epochs with higher likelihood of slow-wave sleep and epochs closer to the end of sleep are assigned higher weights.

### Statistical Analysis

Two datasets were constructed from the sample. A daily dataset restricted to days with an explicit marijuana response supported within-person analysis of nocturnal cardiac autonomic physiology and sleep. A weekly dataset aggregated those days into consecutive seven-day blocks numbered from each member’s first day logging marijuana use/non-use, retaining weeks that contained at least three marijuana responses, and was used to examine use probability over time.

Daily outcomes included RHR, HRV, sleep duration, sleep disturbances, wake time, and time in wearable-estimated sleep stages. Each was analyzed using a linear mixed model with a random intercept for each member. Marijuana use was recorded as a daily binary indicator and partitioned by person-mean centering into a between-person term — an individual’s proportion of logged days with marijuana use, ranging from 0 to 1 — and a within-person term capturing daily deviations from that proportion. Between-person estimates represent the contrast between use on all logged days and use on none. Models were adjusted for gender, age at activation, height, weight, weekend versus weekday, season, device generation, and sleep-onset timing. Gender effects were assessed by adding (within- and between-person) marijuana use interaction terms to the primary models, estimating gender associations as simple slopes. In sensitivity analyses, models were adjusted for self-reported alcohol use, and separately, previous day physical activity, each decomposed into between- and within-person terms.

Weekly marijuana use was modelled using binomial generalized linear mixed models with a logit link, with the counts of use and non-use days within each person-week as the two-column response. To reduce the influence of week-to-week sampling variability and limit multiple comparisons, weeks were collapsed into six 12-week quarters. Models were adjusted for age at activation, gender, season, the number of weeks each member contributed, and the proportion of each week’s logged days falling on a weekend. Marginal predicted probabilities of daily use were estimated for each quarter. Change from baseline was assessed by comparing each subsequent quarter with the first on the probability scale (Dunnett-adjusted), and quarter-to-quarter change by contrasting each consecutive quarter, adjusted for multiplicity (multivariate-t). Effect moderation was assessed by adding interactions between quarter and gender, age group (18–39, 40–59, and 60–79 years), and tertile of first-quarter use probability (i.e., low, medium, and high), with between-group differences in the change from the first to the sixth quarter tested on the log-odds scale. Sensitivity analyses restricted the sample to members who contributed data in all six quarters and refit the full-sample model without the covariate for the total number of weeks each member contributed. We also adjusted the trajectory model for weekly physical activity, decomposed into between- and within-person terms to test whether the decline reflected concurrent changes in activity. As a further sensitivity analysis, quarterly use was re-estimated with inverse-probability-of-censoring weights to account for differential attrition, using a pooled logistic model of continued observation adjusted for quarter, age at activation, gender, BMI, and quarterly use probability.

Means are reported ±SD or with 99.5% CIs. Significance was set against a pre-specified α of 0.005. Analyses were conducted in R 4.6.0, using lme4 for linear mixed models, glmmTMB for binomial mixed models, and emmeans for marginal means and contrasts.

## RESULTS

### Sample Characteristics

Our study consisted of 16,965 individuals who contributed 5,226,739 person-days of data, and 819,963 person-weeks to trajectory models (**Table 1**). Members reported marijuana use on a median of 25.1% of logged days (IQR, 7.1–62.6%). Daily sensitivity models were fit to 16,027 members (4,710,723 person-days) for the alcohol analysis and 16,965 members (4,791,066 person-days) for the physical activity analysis. Weekly sensitivity models used the full sample of 819,963 person-weeks, except the balanced-panel analysis, which was restricted to 7,367 members contributing 475,014 person-weeks.

**Table 1.** Demographic, cardiac autonomic, and sleep characteristics summarized across individuals marijuana users. All cardiac autonomic and sleep variables are summarized across all available person-days (n = 5,222,356 for cardio-autonomic, n = 5,226,739 for sleep). Values are unadjusted means and reported as mean ± SD.

| Characteristic | n = 16,595 |
| --- | --- |
| **Demographics** |  |
| Gender (M/W) | 10,000/6,965 |
| Age (years) | 31.9 (9.1) |
| Height (m) | 1.75 (0.10) |
| Weight (kg) | 77.8 (16.0) |
| **Cardiac Autonomic** |  |
| Resting Heart Rate (bpm) | 59.7 (9.6) |
| Heart Rate Variability (ms) | 60.0 (31.2) |
| **Sleep** |  |
| Sleep Duration (min) | 427.1 (83.2) |
| Disturbances (night<sup>-1</sup>) | 11.7 (5.2) |
| Arousal Time (min) | 24.1 (11.7) |
| Wake Time (min) | 53.6 (34.0) |
| Sleep Efficiency (%) | 89.6 (6.1) |
| REM Sleep (%) | 23.8 (9.0) |
| REM Sleep (min) | 101.9 (43.7) |
| Slow-wave Sleep (%) | 21.3 (5.8) |
| Slow-wave Sleep (min) | 90.3 (27.9) |

### Nocturnal Cardiac Autonomic Physiology

On days where marijuana use was reported, an individual’s RHR (-0.12bpm [-0.14, -0.10]; p<0.001) and HRV (-0.47ms [-0.53, -0.42]; p<0.001) were lower than typical (**Figure 1**). At the between-person level, marijuana use was not associated with RHR (0.42bpm [-0.06, 0.90]; p=0.015) or HRV (1.45ms [-0.19, 3.08]; p=0.013).

**Figure 1:**
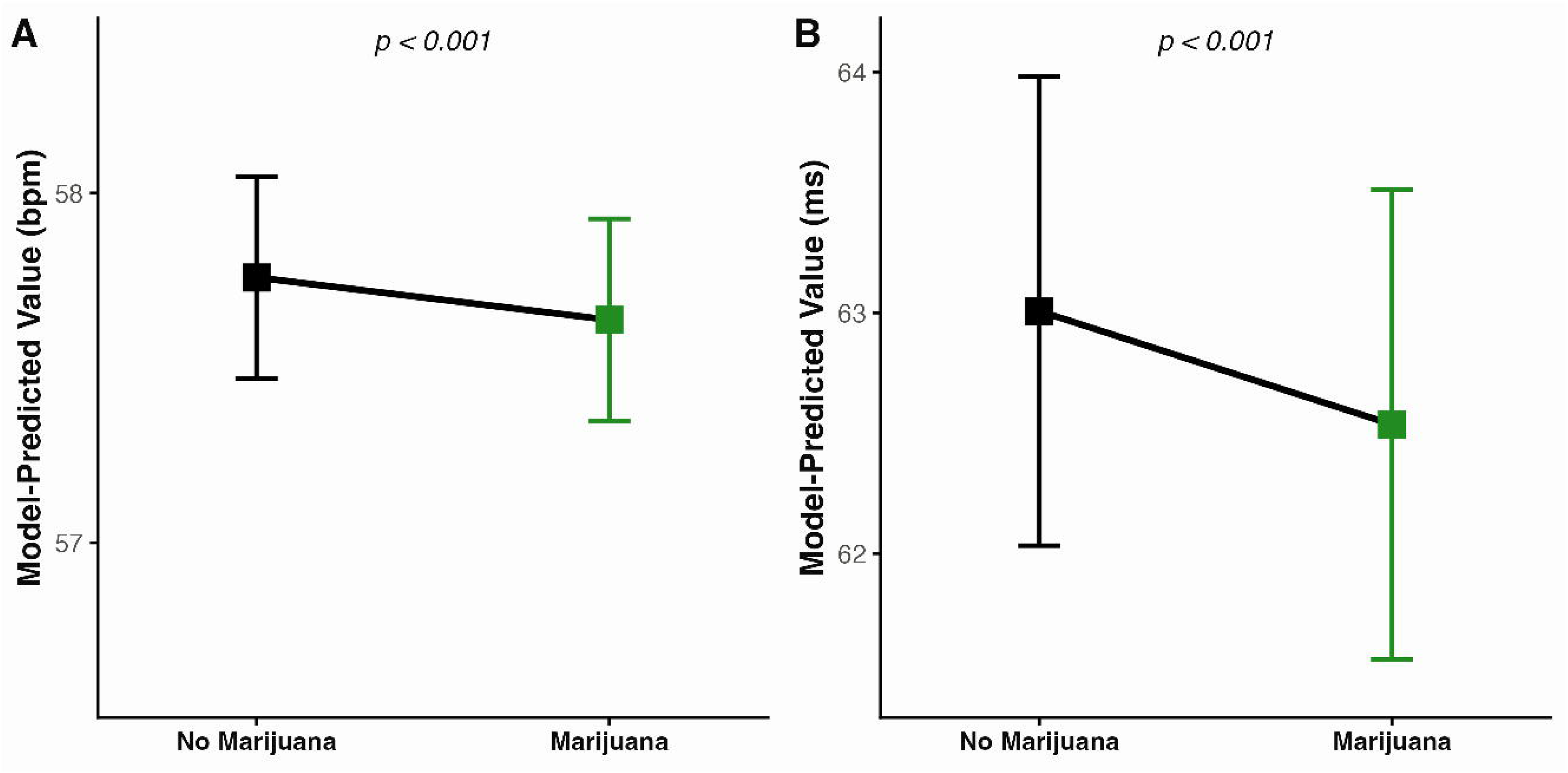
Model-predicted resting heart rate (A) and heart rate variability (B) on a non-use (”No Marijuana”) versus use (”Marijuana”) night, in native units. Estimates are from daily linear mixed models (n = 16,965 individuals; 5,222,356 person-nights) in which marijuana exposure is modeled as a within-person component, representing a night’s deviation from the member’s own average use. Models adjust for gender, age, height, weight, weekday/weekend, season, device model, and bedtime (within-person deviation and person mean). Symbols represent model-predicted values and error bars are 99.5% Wald confidence intervals.

Sensitivity models largely confirmed these findings (**Supplementary Table 1**). Within-person associations were consistent in direction and significance across both models. At the between-person level, RHR remained unassociated with marijuana use after adjustment for alcohol use, but higher habitual marijuana use was associated with elevated RHR after adjustment for physical activity. The lack of between-person associations in HRV persisted in both models. In models including marijuana-by-gender interaction terms, the within-person association between marijuana use and RHR was observed in women (−0.27bpm [−0.30, −0.24]; p<0.001) but not in men (−0.02bpm [−0.05, 0.00]; p=0.009). Conversely, the within-person association between marijuana use and HRV was observed in men (−0.78ms [−0.85, −0.71]; p<0.001) but not in women (0.01ms [−0.08, 0.09]; p=0.86). At the between-person level, marijuana use was not associated with RHR (0.73bpm [−0.03, 1.49]; p=0.007) or HRV (−0.77ms [−3.33, 1.79]; p=0.40) in women. In men, greater marijuana use was not associated with RHR (0.21bpm [−0.42, 0.84]; p=0.35) but was associated with higher HRV (2.95ms [0.84, 5.07]; p<0.001).

### Sleep Fragmentation and Architecture

Marijuana use was associated with differences in sleep duration and fragmentation at both the within- and between-person level (**Figure 2**). Individuals tended to sleep longer than typical following marijuana use days (7.60min [7.37, 7.83]; p<0.001). However, sleep following marijuana use was more fragmented, with more disturbances (+0.47 [0.45, 0.48]; p<0.001), more wake and (1.79min [1.70, 1.89]; p<0.001) arousal (1.04min [1.01, 1.08]; p<0.001) time, and lower sleep efficiency (-0.12% [-0.14, -0.10]; p<0.001). At the between-person level, greater habitual marijuana use was associated with shorter sleep duration (-7.70min [-9.98, -5.43]; p<0.001), more disturbances (+0.66 [0.47, 0.85]; p<0.001), greater wake (7.88min [6.68, 9.07]; p<0.001) and arousal time (1.46min [1.03, 1.90]; p<0.001), and lower sleep efficiency (-1.54% [-1.77, -1.31]; p<0.001).

**Figure 2:**
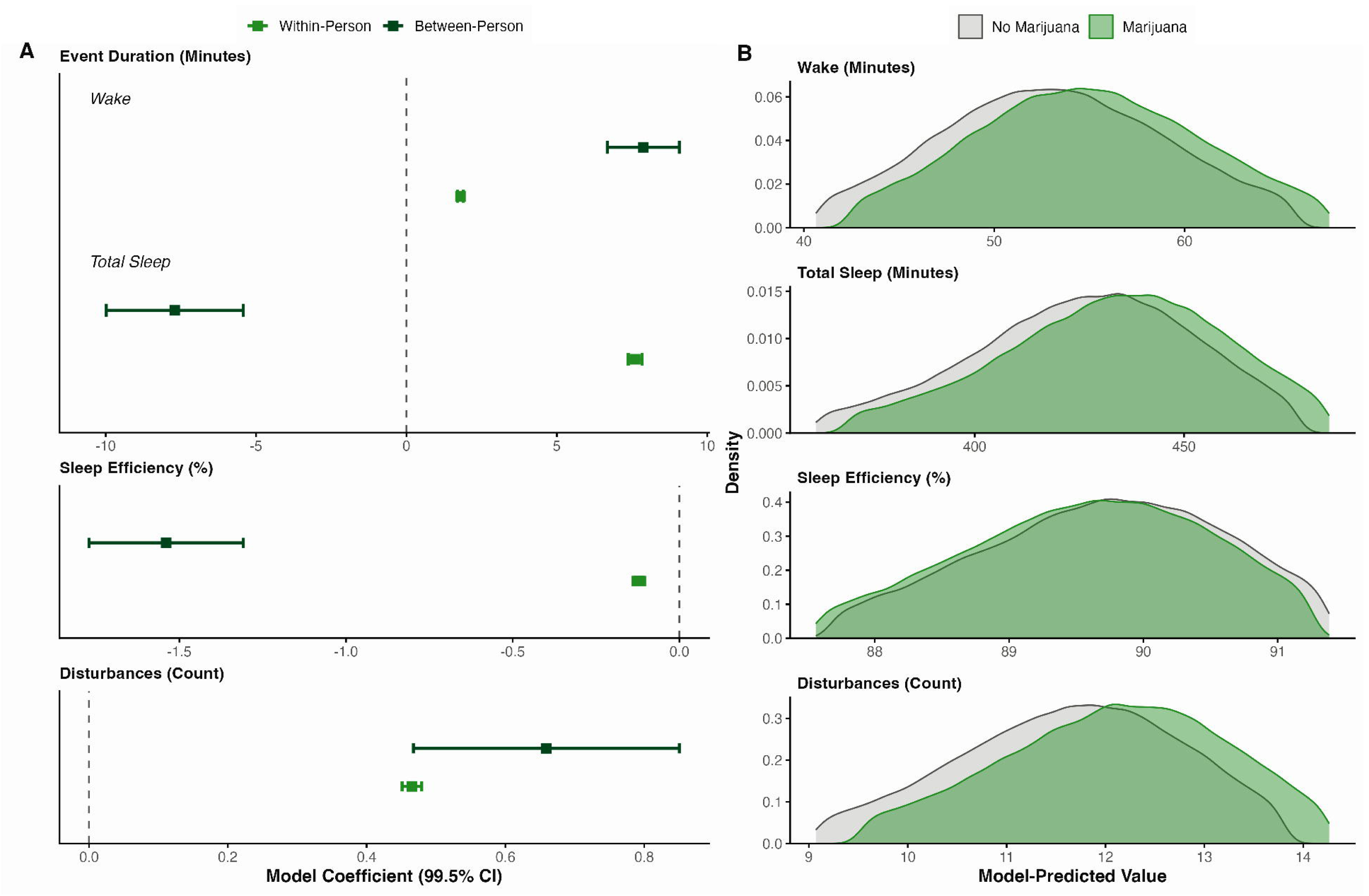
(A) Estimates from daily linear mixed models (n=16,965 individuals, 5,226,739 person-nights) in which marijuana exposure is decomposed into a within-person component representing a night’s deviation from the member’s own average use and a between-person component representing a member’s average use. Models adjust for gender, age, height, weight, weekday/weekend, season, device model, and bedtime (within-person deviation and person mean). Within- and between-person coefficients for sleep measures, grouped by unit type. (B) Distributions of model-predicted values (5^th^ to 95^th^ percentile) on a non-use versus use night for the same outcomes. Symbols represent model coefficients and error bars are 99.5% Wald confidence intervals.

Wearable-estimated sleep architecture also differed with marijuana use. Marijuana-use days were characterized by less REM sleep, both as a percentage of total sleep (-0.85% [-0.87, -0.82]; p<0.001) and in minutes (-1.84min [-1.96, -1.72]; p<0.001). Slow-wave sleep was also lowered both as a percentage of total sleep (-0.50% [-0.51, -0.48]; p<0.001) and in minutes (-0.45min [-0.53, -0.37]; p<0.001). At the between-person level, higher marijuana use was associated with lower REM (% Total Sleep: -1.43% [-1.80, -1.07]; p<0.001; Duration: -7.74min [-9.39, -6.09]; p<0.001) and slow-wave sleep (%Total Sleep: -1.24% [-1.46, -1.03]; p<0.001; Duration: -6.95 [-7.92, -5.97]; p<0.001).

All within- and between-person associations with sleep outcomes were unchanged in both sensitivity models (**Supplementary Table 1**). Similarly, sleep outcomes were consistent across gender, with the within- and between-person associations with marijuana use remaining significant and direction concordant (**Supplementary Table 2**).

### Marijuana Use Trajectories

The model-predicted probability of marijuana use declined across the 72-week observation period (**Figure 3A**), with all quarters demonstrating lower probabilities than the first, and significant declines in each consecutive quarter (all p<0.001). Trajectories differed by baseline use tertile (**Figure 3B**). Use increased in the lowest tertile in all consecutive quarters except from quarter 4 to quarter 5 (p=0.015). Use declined in the middle tertile in each consecutive quarter (all p<0.001), with the exception of quarter 5 to quarter 6 (p=0.97). Use declined in the highest tertile in each consecutive quarter (all p<0.001). The highest use tertile declined to a significantly greater degree than did the middle tertile (p<0.001), as did the middle to the low (p<0.001). Use declined significantly in both genders (**Figure 3C**) in all consecutive quarters (all p<0.001) but to a greater degree in men (p<0.001). Use declined across each age group (**Figure 3D**), with the magnitude of decline increasing in a stepwise fashion from young to old (all p<0.001). The overall decline was unchanged across sensitivity analyses restricting to members present in all six quarters, removing the total number of weeks contributed as a covariate, adjusting for weekly physical activity, and weighting for differential attrition (**Supplementary Table 3**).

**Figure 3:**
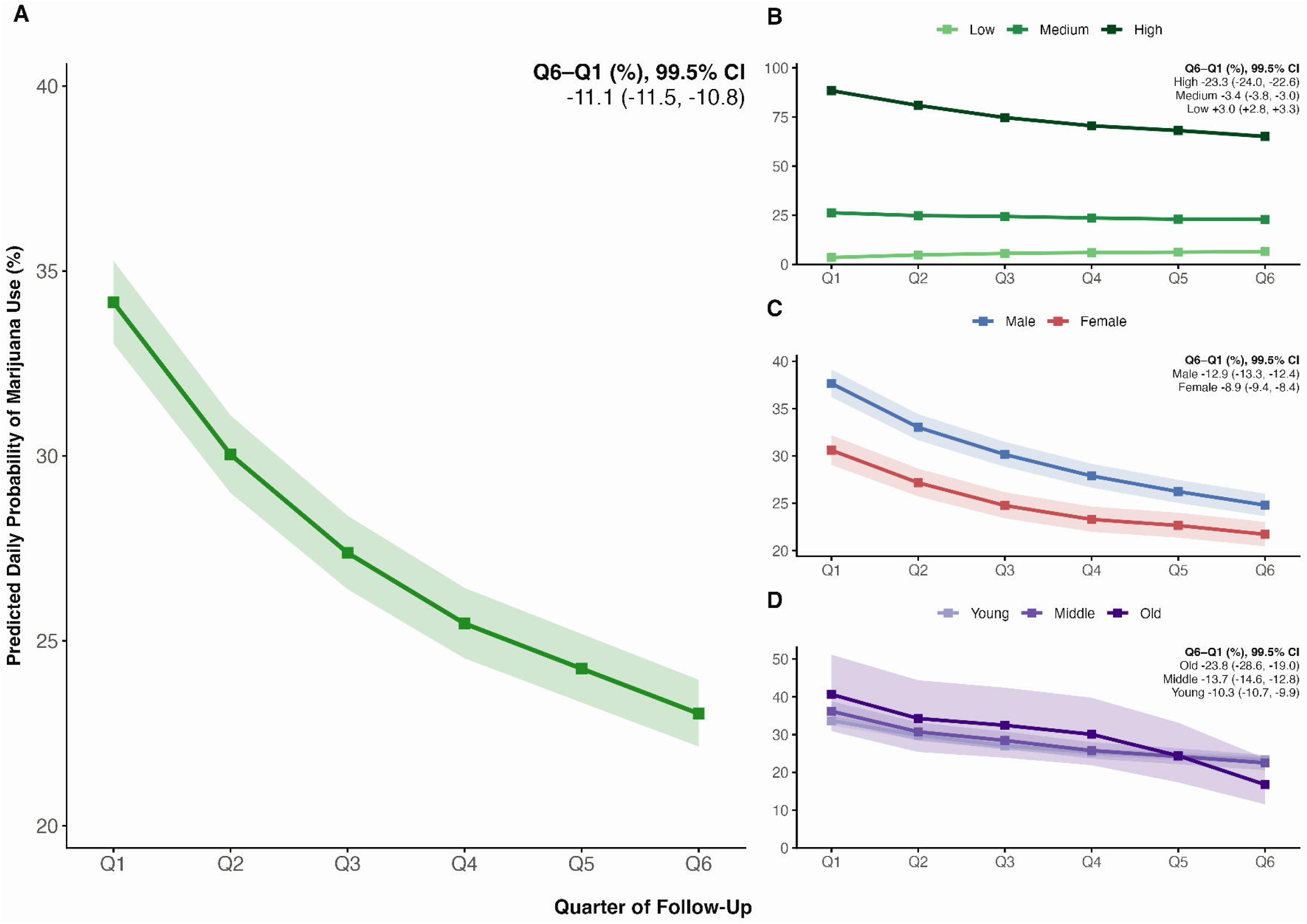
(A) Model-predicted daily probability of marijuana use by quarter over the 504 day window after a user’s first marijuana log from generalized linear mixed models fit to 819,963 user-weeks from 16,965 members. (B) Estimates form models stratified by marijuana use tertile, (C) gender, and (D) age tertile. Insets give the Q6−Q1 change in predicted probability (% points) with 99.5% CIs. Symbols represent model-predicted probabilities and shaded bands are 99.5% confidence intervals.

## DISCUSSION

In the present study, we used large scale, real-world data to understand how marijuana use associates with nocturnal cardiac autonomic physiology, sleep, and longitudinal use patterns. Within individuals, marijuana use days were characterized by lower nocturnal RHR and HRV and longer but more disrupted sleep, including changes in wearable-estimated sleep architecture. More frequent habitual use was also associated with greater sleep fragmentation. Over time, marijuana use declined, with the largest declines among those reporting the most frequent initial use. These findings demonstrate measurable physiological and sleep differences associated with marijuana use in everyday life and suggest that longitudinal tracking may provide insight into changes in use behavior. In doing so, this study constitutes, to our knowledge, the largest and most comprehensive characterization of the physiological and behavioral correlates of habitual marijuana use in a free-living population.

Marijuana’s effects on cardiac autonomic function are a growing interest, particularly given studies linking use to adverse cardiovascular outcomes (Jeffers et al., 2024; Kamel et al., 2025). Laboratory studies administering marijuana have identified robust effects, including a dose-dependent tachycardia attenuated by beta-adrenergic blockade with propranolol (Renault et al., 1971; Sulkowski et al., 1977), alongside reduced vagal control of heart rate and lower HRV during wakefulness (Nardone et al., 2023). Recently a placebo-controlled trial extended these reductions in vagally mediated HRV to the sleep period (Gonzalez et al., 2026). Our within-person finding of a lower RHR on use days, observed in women but not men, runs counter to the acute tachycardia observed in laboratory settings. This discrepancy likely reflects differences in measurement timing. Acute heart-rate increases persist for only several hours (Spindle et al., 2018), and oral THC administered before bedtime elevates heart rate during sleep, but predominantly across the first ∼6 hours of the night when THC concentrations are highest, resolving toward morning (Gonzalez et al., 2026). Our RHR, drawn from the final hours of sleep, likely falls after the acute tachycardia has subsided, reconciling our lower nocturnal values with reports of transient early-night elevations. In contrast, lower HRV on use days, observed in men but not women, was consistent with laboratory evidence and may reflect effects on cardiac autonomic control independent of heart rate. These gender-specific patterns should be interpreted cautiously, as controlled human laboratory studies have found only inconsistent evidence of such differences in the acute effects of cannabis (Wright et al., 2026). They may also reflect behavioural rather than physiological differences, as marijuana use in this cohort was self-selected and unmeasured in dose, and women have been shown to self-titrate to lower amounts of cannabis than men, achieving lower THC exposure while experiencing comparable acute effects (Matheson et al., 2020). Collectively, our findings suggest that marijuana use is associated with small but measurable changes in nocturnal cardiac autonomic physiology, and are more apparent within individuals following use than across individuals with differing habitual use.

Sleep is among the most frequently cited reasons for marijuana use, with users describing it as a sleep aid (Dzierzewski et al., 2026). Consistent with that perception, individuals slept longer on marijuana use days. This finding is particularly notable since controlled laboratory experiments have generally not found that cannabis lengthens sleep (Velzeboer et al., 2025), although early experiments did report increased sleep duration with marijuana exposure (Pivik et al., 1972). Conversely, chronic users have been reported to sleep less (Bolla et al., 2008; Gonzalez et al., 2026), consistent with our finding that sleep duration is shorter with greater habitual use. This association should be interpreted cautiously, however, as marijuana withdrawal may itself reduce sleep duration (Feinberg et al., 1976, 1975), or heavier users may use more because they have greater difficulty with sleep themselves. Importantly, the longer sleep observed on marijuana use nights was less continuous, showing more disturbances, more time awake, and lower efficiency. These findings align with the largest polysomnographic study to date examining sleep in marijuana users and non-users, which reported higher wake after sleep onset, lower sleep efficiency, elevated light-stage sleep, and shorter sleep duration in users (Velzeboer et al., 2026). Our findings extend this evidence to free-living conditions and demonstrate that differences in sleep fragmentation occur within individuals across marijuana use and non-use days, rather than only between users and non-users.

Our analyses also indicated less wearable-derived REM sleep and slow-wave sleep. Earlier investigations have also reported slow-wave sleep and REM sleep suppression (Feinberg et al., 1976, 1975; Freemon, 1974; Tassinari et al., 1976). However, a recent meta-analysis found no consistent effect of marijuana administration on REM sleep or slow-wave sleep, and attributed these earlier findings to very high administered doses and enrollment of only small numbers of often marijuana-naive participants (Velzeboer et al., 2025). Thus, our findings align with earlier administration studies but differ from much of the contemporary literature. Given the limitations of sleep staging using accelerometry and photoplethysmography (Birrer et al., 2024), these findings should be interpreted cautiously and motivate further study of sleep architecture during habitual, real-world marijuana use.

We observed a progressive decline in marijuana use across 72 weeks of monitoring, regardless of age or gender and robust to logging frequency and attrition. This pattern persisted when analyses were restricted to participants contributing data across all six quarters and when adjustment for total weeks contributed was removed, and parallels prior reports that tobacco and alcohol use decline during longitudinal wearable monitoring (Curran et al., 2026; G. J. Grosicki et al., 2026). To our knowledge, this is the first evidence that such a pattern extends to marijuana use. Repeated self-monitoring, physiological feedback, or both could plausibly contribute to declining use, consistent with evidence that physiological feedback can change substance use behavior (Eddie et al., 2025). However, our observational design cannot establish causality. Use declined most among participants with the highest initial use, a group of particular concern given rising rates of cannabis use disorder (Han et al., 2026), although this finding warrants particular caution because more frequent users are more likely to regress to the mean. Consistent with that possibility, the lowest users increased their marijuana consumption across the course of the study. Prospective studies are needed to determine whether wearable-based monitoring or feedback can actively reduce marijuana use.

Several limitations should be considered. First, the observational design precludes causal inference, and although within-person modeling controls for stable characteristics, unmeasured time-varying factors may influence both use and outcomes. Second, use was self-reported and binary, so the influence dose, method of exposure, primary cannabinoid exposure, and timing are not captured. Third, sleep and autonomic metrics were wearable-derived rather than polysomnographic, so our REM and slow-wave findings warrant more caution than our duration and fragmentation measures. Fourth, the baseline-use tertiles were defined by first-quarter use while the outcome was change from that quarter, so the extreme strata are expected to regress toward the mean and should be interpreted cautiously. Finally, participants elected to track marijuana use and may differ systematically from marijuana users who do not engage in self-monitoring, limiting generalizability.

In this large, free-living cohort, marijuana use days were characterized by lower nocturnal RHR and HRV and longer but more fragmented sleep, including less wearable-estimated REM sleep and slow-wave sleep. In more frequent users, sleep was similarly fragmented, but also shorter. Finally, use declined steadily over the 72-week window, most steeply among the heaviest users, supporting the potential for such devices to support reducing marijuana use. Together, these findings extend controlled-laboratory evidence to habitual marijuana use in everyday life and demonstrate the potential of consumer wearables to characterize its physiological, sleep, and behavioral correlates at scale. As marijuana increasingly becomes accessible and commonly used, these findings challenge perceptions of uniformly beneficial sleep effects and support further investigation of wearable-based approaches to monitor marijuana use.

## CONTRIBUTORS

DC, GJG, and CPC conceived and designed the study, with contributions from WvH, FF, SRJ, JL, and KEH. DC, GJG, and CPC led the methodology. DC, GJG and CPC led data curation and formal analysis. CPC prepared all figures. CPC drafted the manuscript. All authors reviewed and edited the manuscript and approved the final version.

## DATA AVAILABILITY

The participant data are not publicly available due to intellectual property concerns of WHOOP, Inc. Deidentified participant data and a data dictionary may be made available upon reasonable request to WHOOP (via). Access requires submission of a methodologically sound proposal, approved by WHOOP’s research team, and a signed data use agreement. Requests will receive a response within 4 weeks.

## DECLARATION OF GENERATIVE AI IN SCIENTIFIC WRITING

The authors used the generative AI tool Claude (Anthropic) for language editing and code review. After using this tool, the authors reviewed and edited the content as needed and take full responsibility for the content of the published article.

---

**Supplementary Table 1.** Within- and between-person associations of daily marijuana use with each nocturnal cardiac autonomic and sleep outcome, estimated from linear mixed models with a per-member random intercept in which marijuana use was partitioned into a between-person term (each member’s mean proportion of use days) and a within-person term (each day’s deviation from that mean). Between-person estimates represent the contrast between use on all logged days and use on none. Models were adjusted for gender, age at activation, height, weight, weekend versus weekday, season, device generation, and sleep-onset timing. Each model was additionally adjusted for self-reported alcohol use and for previous-day physical activity, each decomposed into between- and within-person terms in the same manner as marijuana use. Data are estimates (99.5% CI) in each outcome’s native units.

| Outcome | + Alcohol | p | + Physical Activity | p |
| --- | --- | --- | --- | --- |
| **Within Person** |  |  |  |  |
| Resting Heart Rate (bpm) | -0.22 (-0.24, -0.20) | <0.001 | -0.17 (-0.19, -0.15) | <0.001 |
| HRV (ms) | -0.31 (-0.36, -0.25) | <0.001 | -0.38 (-0.44, -0.32) | <0.001 |
| Wake duration (min) | 1.73 (1.63, 1.83) | <0.001 | 1.81 (1.71, 1.91) | <0.001 |
| Total sleep (min) | 7.88 (7.64, 8.13) | <0.001 | 7.84 (7.61, 8.08) | <0.001 |
| Sleep efficiency (%) | -0.10 (-0.12, -10.08) | <0.001 | -0.12 (-0.14, -0.10) | <0.001 |
| Disturbances (count) | 0.47 (0.46, 0.49) | <0.001 | 0.48 (0.46, 0.49) | <0.001 |
| Arousal time (min) | 1.08 (1.04, 1.11) | <0.001 | 1.08 (1.05, 1.12) | <0.001 |
| REM sleep (%) | -0.84 (-0.86, -0.81) | <0.001 | -0.84 (-0.86, -0.81) | <0.001 |
| REM duration (min) | -1.73 (-1.86, -1.60) | <0.001 | -1.74 (-1.86, -1.61) | <0.001 |
| Slow-wave sleep (%) | -0.51 (-0.53, -0.50) | <0.001 | -0.51 (-0.52, -0.49) | <0.001 |
| Slow-wave duration (min) | -0.44 (-0.52, -0.35) | <0.001 | -0.43 (-0.52, -0.35) | <0.001 |
| **Between Person** |  |  |  |  |
| Resting heart rate (bpm) | 0.45 (-0.04, 0.95) | 0.010 | 1.27 (0.84, 1.71) | <0.001 |
| HRV (ms) | 1.27 (-0.41, 2.95) | 0.034 | -0.98 (-2.50, 0.54) | 0.070 |
| Wake duration (min) | 7.84 (6.61, 9.07) | <0.001 | 7.97 (6.77, 9.18) | <0.001 |
| Total sleep (min) | -7.28 (-9.61, -4.94) | <0.001 | -7.07 (-9.36, -4.79) | <0.001 |
| Sleep efficiency (%) | -1.52 (-1.75, -1.28) | <0.001 | -1.55 (-1.78, -1.31) | <0.001 |
| Disturbances (count) | 0.67 (0.47, 0.87) | <0.001 | 0.65 (0.46, 0.85) | <0.001 |
| Arousal time (min) | 1.52 (1.07, 1.97) | <0.001 | 1.48 (1.04, 1.92) | <0.001 |
| REM sleep (%) | -1.48 (-1.85, -1.10) | <0.001 | -1.45 (-1.82, -1.08) | <0.001 |
| REM duration (min) | -7.84 (-9.55, -6.13) | <0.001 | -7.68 (-9.34, -6.02) | <0.001 |
| Slow-wave sleep (%) | -1.25 (-1.47, -1.03) | <0.001 | -1.14 (-1.35, -0.92) | <0.001 |
| Slow-wave duration (min) | -6.91 (-7.92, -5.91) | <0.001 | -6.37 (-7.34, -5.40) | <0.001 |

**Supplementary Table 2.** Within- and between-person associations of daily marijuana use with each nocturnal sleep outcome, by gender, estimated from linear mixed models with a per-member random intercept in which marijuana use was partitioned into a between-person term (each member’s mean proportion of use days) and a within-person term (each day’s deviation from that mean) and allowed to interact with gender. Between-person estimates represent the contrast between use on all logged days and use on none. Gender-specific estimates are the within- and between-person associations for women and men obtained from the marijuana-by-gender interaction. Models were adjusted for age at activation, height, weight, weekend versus weekday, season, device generation, and sleep-onset timing. Data are estimates (99.5% CI) in each outcome’s native units.

| Outcome | Men | p | Women | p |
| --- | --- | --- | --- | --- |
| **Within Person** |  |  |  |  |
| Wake duration (min) | 1.90 (1.77, 2.02) | <0.001 | 1.64 (1.48, 1.79) | <0.001 |
| Total sleep (min) | 7.83 (7.54, 8.13) | <0.001 | 7.25 (6.89, 7.61) | <0.001 |
| Sleep efficiency (%) | -0.13 (-0.15, -0.11) | <0.001 | -0.10 (-0.13, -0.08) | <0.001 |
| Disturbances (count) | 0.54 (0.52, 0.55) | <0.001 | 0.36 (0.33, 0.38) | <0.001 |
| Arousal time (min) | 1.23 (1.19, 1.27) | <0.001 | 0.76 (0.71, 0.81) | <0.001 |
| REM sleep (%) | -0.87 (-0.90, -0.84) | <0.001 | -0.81 (-0.85, -0.77) | <0.001 |
| REM duration (min) | -1.84 (-2.00, -1.69) | <0.001 | -1.84 (-2.03, -1.65) | <0.001 |
| Slow-wave sleep (%) | -0.52 (-0.54, -0.49) | <0.001 | -0.47 (-0.50, -0.45) | <0.001 |
| Slow-wave duration (min) | -0.39 (-0.49, -0.28) | <0.001 | -0.55 (-0.68, -0.42) | <0.001 |
| **Between Person** |  |  |  |  |
| Wake duration (min) | 6.79 (5.24, 8.34) | <0.001 | 9.47 (7.59, 11.35) | <0.001 |
| Total sleep (min) | -8.56 (-11.52, -5.61) | <0.001 | -6.44 (-10.02, -2.87) | <0.001 |
| Sleep efficiency (%) | -1.39 (-1.69, -1.09) | <0.001 | -1.76 (-2.12, -1.39) | <0.001 |
| Disturbances (count) | 0.58 (0.34, 0.83) | <0.001 | 0.77 (0.47, 1.07) | <0.001 |
| Arousal time (min) | 1.33 (0.77, 1.90) | <0.001 | 1.66 (0.97, 2.34) | <0.001 |
| REM sleep (%) | -1.46 (-1.93, -0.99) | <0.001 | -1.40 (-1.97, -0.83) | <0.001 |
| REM duration (min) | -7.94 (-10.08, -5.80) | <0.001 | -7.44 (-10.03, -4.86) | <0.001 |
| Slow-wave sleep (%) | -1.37 (-1.64, -1.09) | <0.001 | -1.06 (-1.39, -0.72) | <0.001 |
| Slow-wave duration (min) | -7.57 (-8.84, -6.31) | <0.001 | -6.03 (-7.56, -4.50) | <0.001 |

**Supplementary Table 3.** Model-predicted daily probability of marijuana use by quarter over the 72-week window, from four sensitivity analyses: restriction to individuals contributing data in all six quarters (Balanced); refitting without the covariate for the total number of weeks each member contributed (− Weeks); additional adjustment for weekly physical strain decomposed into between- and within-person terms (+ Physical Activity); and inverse-probability-of-censoring weighting to correct for differential attrition, estimated from a pooled logistic retention model (quarter, age at activation, gender, BMI, quarterly use proportion) and winsorised at the 1st/99th percentile (IPCW). In every model each quarter differed from the first at p < 0.001 (Dunnett-adjusted). Data are estimates (99.5% CI).

| Quarter | Balanced (%) | - Weeks (%) | + Physical Activity (%) | IPCW (%) |
| --- | --- | --- | --- | --- |
| Q1 | 37.3 (36.1–38.6) | 32.7 (32.0–33.4) | 34.2 (33.4–35.0) | 34.0 (32.9–35.1) |
| Q2 | 33.9 (32.8–35.1) | 28.7 (28.0–29.4) | 30.0 (29.3–30.8) | 29.8 (28.8–30.9) |
| Q3 | 31.8 (30.7–32.9) | 26.1 (25.5–26.7) | 27.4 (26.7–28.1) | 27.2 (26.2–28.2) |
| Q4 | 29.7 (28.6–30.8) | 24.2 (23.6–24.9) | 25.5 (24.8–26.1) | 25.3 (24.3–26.2) |
| Q5 | 28.0 (26.9–29.0) | 23.1 (22.5–23.7) | 24.3 (23.6–24.9) | 24.1 (23.2–25.0) |
| Q6 | 26.4 (25.4–27.4) | 21.9 (21.3–22.5) | 23.1 (22.4–23.7) | 22.8 (21.9–23.7) |

